# IMMUNE RESPONSE TO SARS CoV2 INFECTION BY TLR3, TLR4 AND TLR7 GENE EXPRESSION

**DOI:** 10.1101/2023.05.12.23288889

**Authors:** Veronica L. Martinez Marignac, Gloria S. Oertlin, Jose Luis Favant, Erika Fleischman, Mercedes Salinas, Gaston Marchetti, Zaida Gassali, Silvina M. Richard

## Abstract

Toll-like receptors (TLRs) may be involved both in the initial failure of viral clearance and in the subsequent development of severe clinical manifestations of COVID-19, essentially ARDS (acute respiratory distress syndrome) with fatal respiratory failure. We present the gene expression of TLR 3, 4, and 7 in nasopharyngeal total RNA samples from 150 individuals positive for SARS Cov2 (DET) by molecular techniques of isothermal amplification (Neokit SA) and 152 SARS Cov2 non detectable (ND) ambulatory and hospitalized patients with a non-defined respiratory disease, and we compared with the symptomatology developed by all those patients. We analyzed 4 cohorts: 1-SARS Cov2 genome detected patients with severe to high symptomatology (n=107); 2-SARS Cov2 genome detected patients low to mild symptomatology (n=43); 3-SARS Cov2 genome non detected patients with severe to high symptomatology (n=109); and 4-SARS Cov2 genome non detected patients low to mild symptomatology (n=41).

Our results not only contradict few previous study, it also corrects for sample size bias, showing no significant differences of expression for TLR3, TLR4 and TLR7 between SARS Cov2 DET and ND total cohort of patients (Non Paired T –Test p Value>0.1). When compared severity of symptoms -presence of symptoms from the COVID-19 12 WHO diagnosis symptoms- and gene expression, here we found significant positive correlation between severe symptomatology, and the number of symptoms and death for TLR4 and TLR7 for both DET and ND COVID-19 patients. When the cohort was construct with low/middle and severe symptoms, the Correlation Coefficient showed that expression of TLR4 and TLR7 was significantly amplified in those ND patients with severe symptomatology (*p Value*= 0.00311) as well as for TLR3 in ND low to mild symptoms cohort of patients. We also showed and discussed the results obtained of these genes expression and the sex and age of patients. In summary, our data suggest that although our innate immune system with TLRs contributes to the elimination of viruses, it can also be associated with harm to the host due to persistent inflammation and tissue destruction. We confirmed that principally TLR4 and TLR7 could be involved not only in the pathogenesis of COVID□19 but also in other respiratory diseases with same symptomatology. We agree with previous studies that treatments focus on TLR4 and TLR7 expression in inflammatory respiratory diseases could be a start point against severe symptoms development.

## INTRODUCTION

Toll-like receptors (TLR) are a family of receptors comprising 10 members (TLR1-TLR10), which are expressed in innate immune cells such as macrophages, as well as epithelial and fibroblast cells. O’Neill et al (2013) reported TLRs as involved in the initial failure of viral clearance and Onofrio et al (2020) suggested their participation in the subsequent development of fatal clinical manifestations in severe COVID-19 patients, and essentially in ARDS (acute respiratory distress syndrome) they would be related with fatal respiratory failure. TLRs family activation can be induced by a multitude of pathogen-associated molecular patterns (PAMPs) present in bacteria, viruses, and other foreign organisms (Duran et al, 2014). Briefly, it was suggested that TLRs play an important role in initiating innate immune responses, with the production of inflammatory cytokines, type I IFNs, and other mediators (Bortoloti et al, 2021).

TLRs are expressed on the cell surface or in the endosome compartment, such as TLR-1, -2, - 4, -5, -6, -10 and TLR-3, -7, -8, -9, respectively (O’Neill et al, 2013; Onofrio et al 2020). While TLR3 in the endosome compartment recognizes viral double-stranded RNA (dsRNA), TLR7 recognizes viral single-stranded RNA and is therefore suggested to likely be involved in the clearance of SARS-CoV-2 and similar virus (Imai et al, 2008; Onofrio et al 2020). In addition, other studies have shown interactions between protein spike glycoprotein (S) of SARS-CoV2 and the TLR innate immune system, including C-lectin-like receptors (CLR), neuropilin-1 (NRP1), and regulated protein 78 by no immune system receptor for glucose (GRP78) (Gadanec et al, 2021).

An intact SARS-CoV-2 virus is surrounded by a lipid envelope that contains the envelope protein (E), membrane protein (M), and spike glycoprotein (S). The genome of SARS-CoV-2 consists of large, single-stranded positive RNA (from 29.8 to 29.9 kb) that contains 14 open-reading frames (ORFs) encoding 27 proteins and we suggest that in nude could be detected by TLR7. The genome sequence of SARS-CoV-2 displays 79.0% homology with SARS-CoV and 51.8% with MERS-CoV. Nucleocapsid (N) proteins form complexes with genomic RNA for genome packaging, before packaging we suggest that TLR7 and TLR3 could be molecules that could recognizes viral single and double RNA structures.

During viral entry into host cells, the surface trimeric S glycoprotein mediates receptor recognition and viral-host cell membrane fusion. The host protease furin cleaves the S protein into S1 and S2 subunits for preactivation, and the receptor-binding domain (RBD) of S1 binds to angiotensin-converting enzyme 2 (ACE2) expressed on the surfaces of host cells. Then, SARS-CoV-2 enters host cells by either direct fusion or endocytosis (Jung and Lee, 2021).

In case of cell surface expressed TLR4, it has been suggested that it binds SARS-CoV-2 spike glycoprotein and this activates TLR4 signaling to increase cell surface expression of ACE2 facilitating entry of SARS-CoV-2, and destroys the type II alveolar cells that secrete pulmonary surfactants. All these which normally decrease the air/tissue surface tension and negatively feedback by then blocking TLR4 own expression in the lungs what promotes ARDS and inflammation (Manik et al, 2022). Thus, we suggested that an available antagonist against TLR4 or TLR3can prevent the onset of severe COVID-19 or ARS respectively, in symptomatic patients and synergize with active antiviral therapy.

While TLR3 recognizes viral double-stranded RNA (dsRNA), TLR7 recognizes viral single-stranded RNA and is therefore likely to be involved in SARS-CoV-2 clearance. On the other hand, TLR4, at the surface of cells, toll-like receptor 4 (TLR4) in the induction of damaging inflammatory responses during acute viral infections as it functions as a sensor for damage-associated molecular patterns (DAMPs). These include a wide variety of molecules released from injured or dying tissues as well as molecules actively released in response to cellular stress from intact cells.

TLR’s member family also plays an important role in cancer progression. A comparison with cancer treatment is necessary herein, as TLR stimulation of cancer cells can lead to tumor progression or inhibition. Stimulation of TLR 2, 4, and 7/8 can lead to tumor progression through production of immunosuppressive cytokines, increased cell proliferation, and resistance to apoptosis. On the other hand, stimulation of TLR 2, 3, 4, 5, 7/8 and 9, often combined with chemo or immunotherapy, can lead to tumor inhibition through different pathways. In addition, TLR stimulation in NK cells (natural killer cells) and APCs (antigen-presenting DC-dendritic cells and macrophages) can induce CTL (cytotoxic T lymphocytes) to further inhibit tumor growth (Urban-Wojciuk et al, 2019). In consequence, no in cancer treatment but it could be a good idea to activate these pathways in patients who suffered of a severe COVID-19 infection where it could be a disadvantage to develop a cytokine endosplasmatic storm (Chahal et al, 2013; Berzemer and Garzen, 2021). To decrease hyperinflammation and thrombotic complications in vulnerable population with severe stages COVID-19, this could be done by the expression control of TLR3, TLR4 and TLR7 genes, however there is a need of more evidence and highlight the expression level of TLR3, TLR4 and TLR7 (Bortoloti et al, 2021; Manik and Singh, 2021) in ARS and COVID-19 patients.

As the TLRs family members have been studied in the field of oncology, different antitumor treatments have been developed as activators, inhibitors or antagonists of these receptors (Urban Wojciuk, et al 2019). However, there is a few recent published studies showing the gene expression profile of some of this family genes in COVID-19 infected patients (Bagheri-Hosseinabadi et al, 2022; Menezes et al, 2021) on small cohorts of patients, different symptomatology and controls. Furthemore, it also few studies relating treatment of COVID-19 by agonist of TLR7 and TLR8 as for example TLR7 stimulation may not only help viral clearance but also by it anti-inflammatory effect decrease and synergized with severe symptoms treatments (Khalifa and Ghoneim, 2021). As a consequence, we suggest that having a much larger study, with patients of respiratory disease symptoms during the Delta and Omicron waves in Argentina (2020-2021) would decrease biases by number and only one systems of detection of the viral genome. Knowing fully the status of TLR3, TLR4 and TLR7 expression in infected patients with ARS or SARS Cov2, can help in repositioned cancer drugs for ARS treatment. In consequence help focusing studies such as in those presenting recent progress in the development of TLR agonists and antagonists, as immunomodulators including as vaccine adjuvants or agents to treat hyperinflammatory responses during SARS-CoV-2 infection (Dai et al, 2022).

Here, our goal was to evidence TLR 3, 4, and 7 gene expression by semi quantitative retro transcription PCR in nasopharyngeal total RNA samples from patients admitted to the service of 4 public hospitals in Diamante departmental jurisdiction –Diamante, Entre Ríos, Argentina. Due to respiratory infection and symptoms that according to the Health Ministry from Argentina and the WHO interim guidance, were attributed to those for SARS CoV2. We tested the expression of three genes in positive (DET) or negative (ND) for SARS CoV2 infection by molecular isothermal amplification techniques (Neokit, SA, CONICET-Cassara, Argentina). Our data contradict previous study, showing in a larger cohort a no significant TLR3, TLR4 and TLR7 expression differences between non-detected and detected COVID-19. However, our results showed when compared SEVERETY of symptoms and gene expression by a Spearman’s Correlation Coefficient that there was significant positive association between severe symptomatology and death for TLR3 in ND patients, whereas a significant positive correlation to SEVERETY for TLR4 and TLR7 expression for both infected and non COVID-19 infected patients, and the correlation was higher positive with those ND patients. We also show analyses for association between these genes expression, sex and age of patients.

## MATERIALS AND METHODS

### PATIENT COHORTS

Patients were divided into 2 main groups: 1-patients with active SARS-CoV-2 infection confirmed by a new isothermal amplification kit developed by Neokit SA, Argentina, of nasopharyngeal swabs (COVID-19, DET n = 149) and, 2-SARS-CoV-2 negative infected patients with and/or SARS-CoV-2 IgG antibody-negative serology (ND n=152). Disease severity was classified into three clinical types (low/mild/moderate and severe), according to the Health Ministry from Argentina and the WHO interim guidance. We took in account 1=Headache 2=Myalgia 3=Sore throat 4=Fever less than 38ºC 5=Fever>=38 6=cough 7=Rhinitis/Nasal congestion 8=Vomiting 9=Diarrhea 10=respiratory failure 11=anosmia 12=dysgeusia and death.

Low symptomatic patients showed symptoms such as 2, 8 or 9 (n=191), mild/moderate symptomatology patients showed 3, 4 or 5 (n=76) and severe symptomatology was considered with 5, 6, 7, 10, 11 and 12 (n=35).

In brief, patients with low symptoms (n = 191) do not present evidence of viral pneumonia or hypoxia. Symptoms are non-specific and could include separately fatigue, headache, myalgia, abdominal pain, vomiting and diarrhea. Moderate or mild symptoms patients (n = 76) present symptoms and signs of non-severe pneumonia, fever, sore throat (cough or fast breathing) and includes patients with comorbidities such as cardiac or respiratory disease, diabetes. Our cohort of severe symptoms contains patients under severe disease, the majority with fever over 38□C, nasal congestion, respiratory failure, anosmia and dygeusia and 12 patients have been part of those in ND groups (n=8).

The study was conducted in accordance with the Declaration of Helsinki. The Institutional Review Board CEYSTE -Committee of Ethical and Experimental Work Safety (CEYSTE, CONICET) reviewed and approved the informed consent, the sample collection and the overall study protocol reference 00535/2019.

Characteristics of participants with or without COVID-19 are shown in Table I.

**TABLE I.** Characteristics of the cohort of patients with COVID-19 and miscellaneous infection. (N=sample size; DET=detected infection; ND=non detected infection;

| CHARACTERISTICS | COVID-19 DET<br>INFECTION (N= 149) | COVID-19 ND<br>-ARS-INFECTION (N= 152) |
| --- | --- | --- |
| Age/years |  |  |
| <2 Baby | 8 | 19 |
| <7 Child | 21 | 21 |
| < 18 Teens | 17 | 21 |
| < 36 Young adults | 49 | 39 |
| < 65 Adults | 47 | 45 |
| >65 Old Adults | 8 | 7 |
| Sex |  |  |
| Female | 69 | 79 |
| Men | 80 | 73 |
| Symptomatology |  |  |
| Low | 90 | 93 |
| Mild/Moderate | 40 | 33 |
| Severe | 19 | 26 |

### COVID-19 DETERMINATION

We employed isothermal amplification by the COVID-19 NEOKIT-plus TECNOAMI, developed by CONICET and Cassara SA, Argentina. This is a diagnostic test for the simplified molecular detection of the SARS-CoV-2 virus, the etiological agent of atypical pneumonia or severe acute respiratory syndrome. The NEOKIT PLUS system allowed the purification of total mRNA from patients swabs and, according to the manufacturer protocol, we obtained 50 ul of total RNA. We employed 10ul to determine the presence of viral RNA.

### HUMAN cDNA

We characterized the expression of TLR 3, 4, and 7 in nasopharyngeal total RNA samples from 150 individuals positive for COVID-19 by Neokit PLUS SA and 152 non detectable ambulatory and interned patients. Symptomatology, age, and sex were compared using statistical analyses.

We employed 5ul of patients’ total RNA purified by the NEOKIT PLUS system, which were storaged at -80 □C, and a random hexamer retrotranscription 1uM (PB-L SRL, Argentina), dNTPs 500uM and MilliQ water in 15ul final volume performed. This was exposed to thermo-cycling (65°C 5 min and 2 min on ice), and finally, the reaction was carried to a 20ul final volume, by adding 50mM MgCl2, 100U of MMLV, and 4ul of MMLV buffer (PB-L, Argentina) and thermocycling at 25°C for 10 min and 37°C for 50 min.

TLR3, TLR4 and TLR7 GENE EXPRESSION DETERMINATION: 5ul of cDNA were employed in a semi quantitative gene expression protocol. As a housekeeper gene, we employed human B-actin, 200nM of each of the following primers: Forward-GAGCACAGAGCCTCGCCTTT; Reverse-ACATGCCGGAGCCGTTGTC; 1U of Taq polymerase (Pegasus-PB-L SA Argentina); 3mM MgCl2; 200uM dNTPs; thermocycling 94° C 2 min; and 40 cycles of 92° C 45sec, 60° C 45 sec, 72° C 45sec, and an end step of 72° C 5min.

Table II shows the TLRs primers employed, the conditions of end point PCR were for TLR3 and 4 the following: 2mM MgCl2, 200uM dNTPs, 200nM primers and 1U of Taq polymerase (Pegasus –PB-l SA, Argentina). TLR7 was characterized by the following conditions: 2.6mM MgCl2, 200uM dNTPs, 200nM primers and 1U Taq polymerase. The thermocycling conditions for all the genes were the followings: 94° C 2 minutes, followed by 40 cycles of 95° C 30sec, 62° C 1minute, 72°C 1 minute and an end step of 72° C for 5 minutes. To separate PCR products 10μl of each sample was resolved on a 1.2% agarose gel (Sigma) and electrophoresis was performed with 1x TAE buffer (Invitrogen) and a voltage of 95V for 30-40 min. The bands were visualized by using an ultraviolet trans-illumination and digital images were captured by Gel documentary machine (Care stream, Berlin, Germany).

**Table II.**
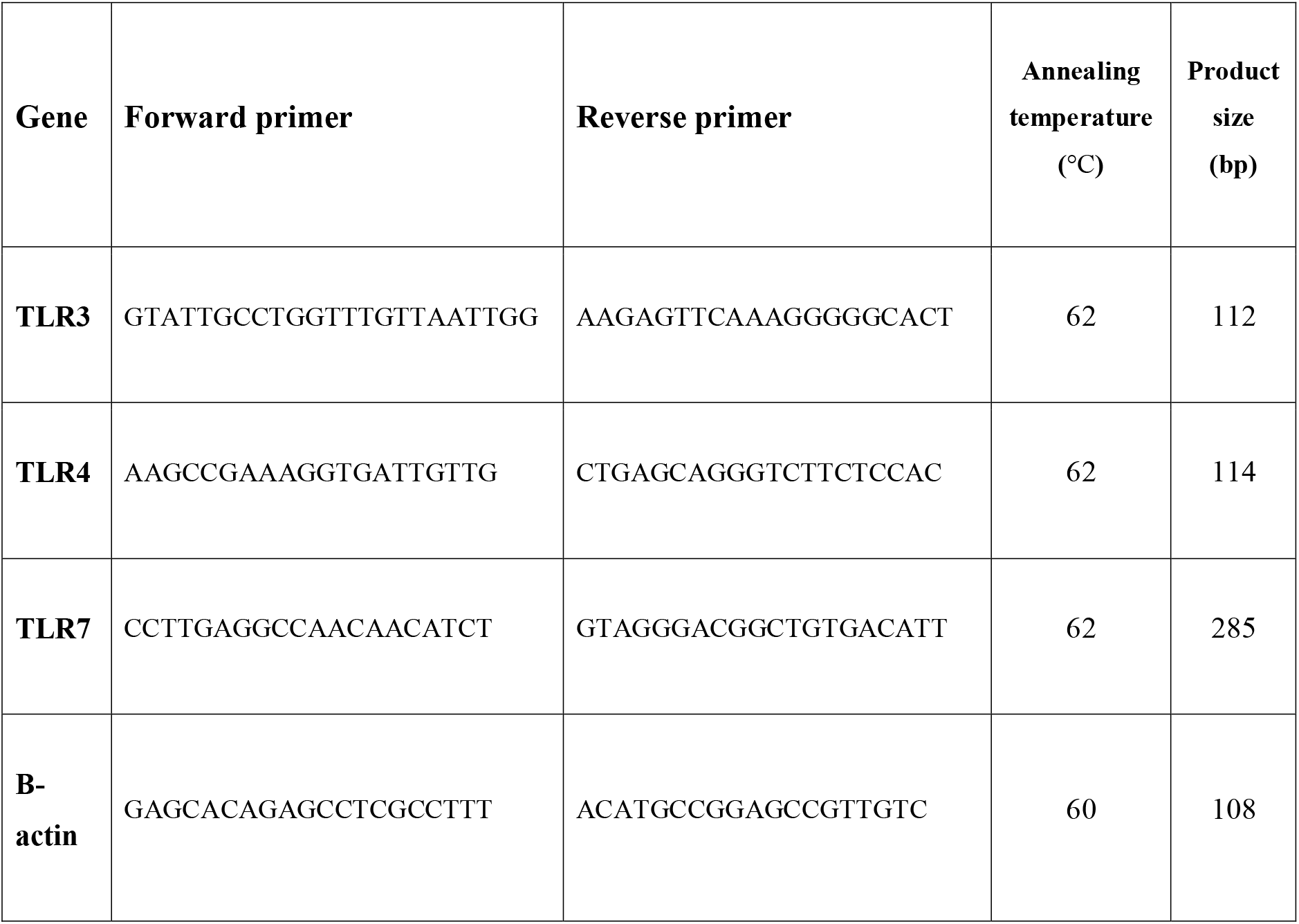
List of TLRs primer sequences used in the experiment for the amplification of human TLR3, 4 and 7 mRNA with RT-PCR (°C=degree Celsius; bp= base pairs).

### STATISTICAL ANALYSIS

For groups and subgroups comparisons we employed a 3nested analysis, non-paired T-Test and Spearman’s Correlation Coefficient to evidence association between level of symptomatology and the expression of these 3 TLRs genes. We also employed a Point-Biserial Correlation Calculator to evidence any correlation between two variables in the special circumstance that one of our variables was dichotomous – DET or ND COVID-19; female or masculine-; we have only two possible values, 1 or 0 for the purposes of this calculator. We employed also a McNemar’s test. We essayed a Welch’s ANOVA for age and sex differences and gene expression. We employed a significant p-Value of 0.05.

## RESULTS

The expression of TLR3, TLR4 and TLR7 genes have no correlation with the presence of COVID-19 viral genome in the total cohort patient sample using the Point-Biserial Correlation calculator (Table 3 and histogram 1). The McNemar’s test did not evidence significant association between severe cases and COVID-19 presence in the patients.

**Histogram 1:**
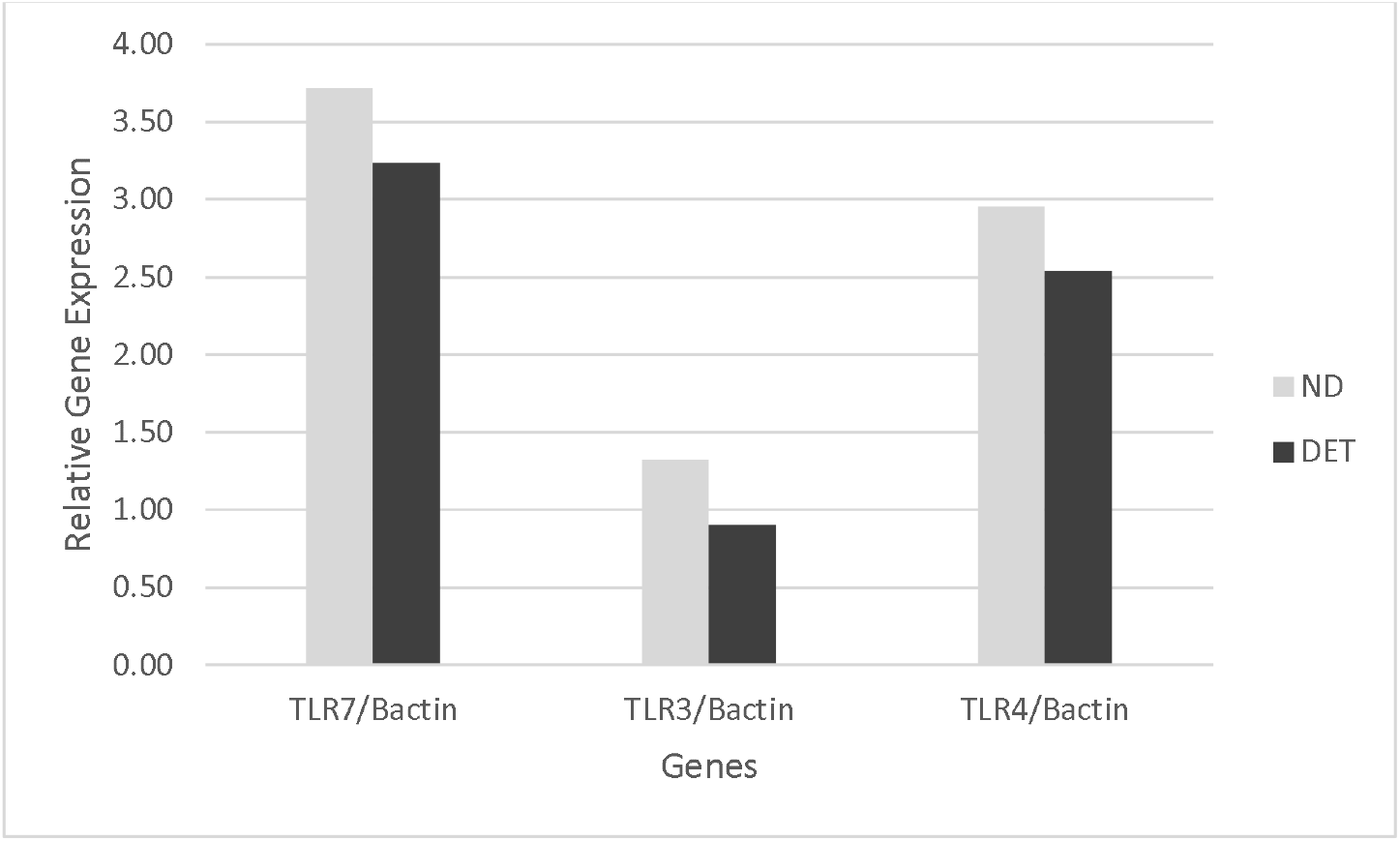
Gene expression compared by ND and DET patients.

**Table 3.** Semi quantitative expression of TLR3, TLR4 and TLR7 corrected by B-actin gene expression.

| ND | TLR7/Bactin | TLR3/Bactin | TLR4/Bactin |
| --- | --- | --- | --- |
| Average | 3,72 | 1,32 | 2,95 |
| SE | 0,02 | 0,01 | 0,02 |

| DET | TLR7/Bactin | TLR3/Bactin | TLR4/Bactin |
| --- | --- | --- | --- |
| Average | 3,24 | 0,90 | 2,54 |
| SE | 0,02 | 0,01 | 0,01 |

By applying Non Paired T-test, we evidenced non-significant differences of expression for TLR3, TLR4 and TLR7 between COVID-19 detected and non-detected patients.

Our results showed non-significant differences of expression for TLR3, TLR4 and TLR7 between SARS Cov2 detected and non-detected patients (Non Paired T –Test p Value>0.1).

On average, all Covid19 detected patients expressed all the studied genes in a same manner as non-detected patients; however, when we accounted for the SEVERITY of symptoms (Table 4 and Histogram 2), we did not evidence correlation between expression and SEVERITY for TLR3 expression in the total sample. Instead, TLR3 expression was significant positive correlated with high SEVERITY (Pearson R= 0.1116, p Value of 0.0519) in ND.

**Table 4.**
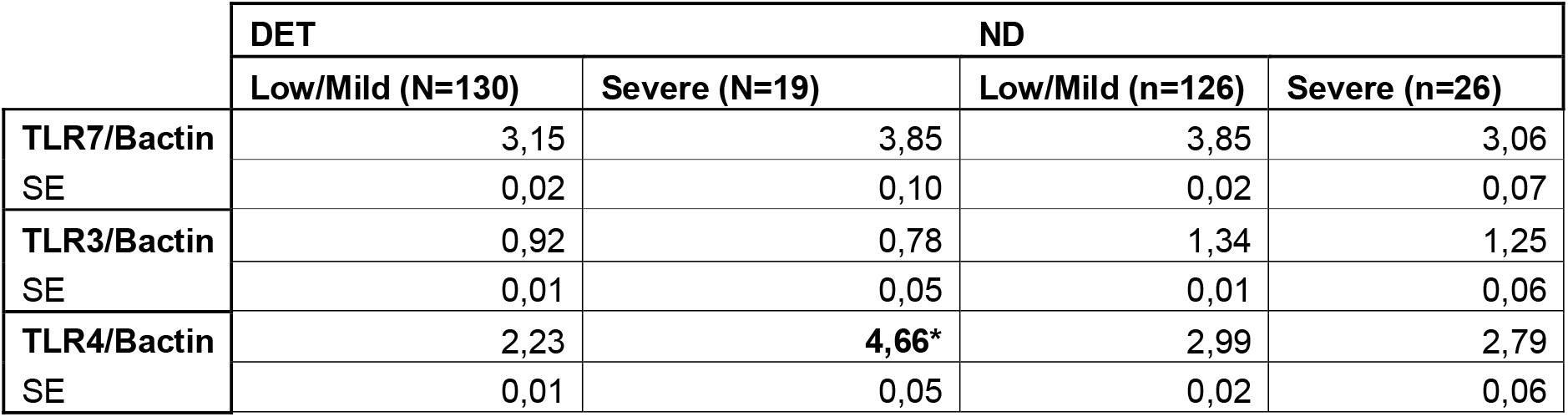
The 4 cohort study and their TLRs semi quantitative gene expression.

**Histogram 2:**
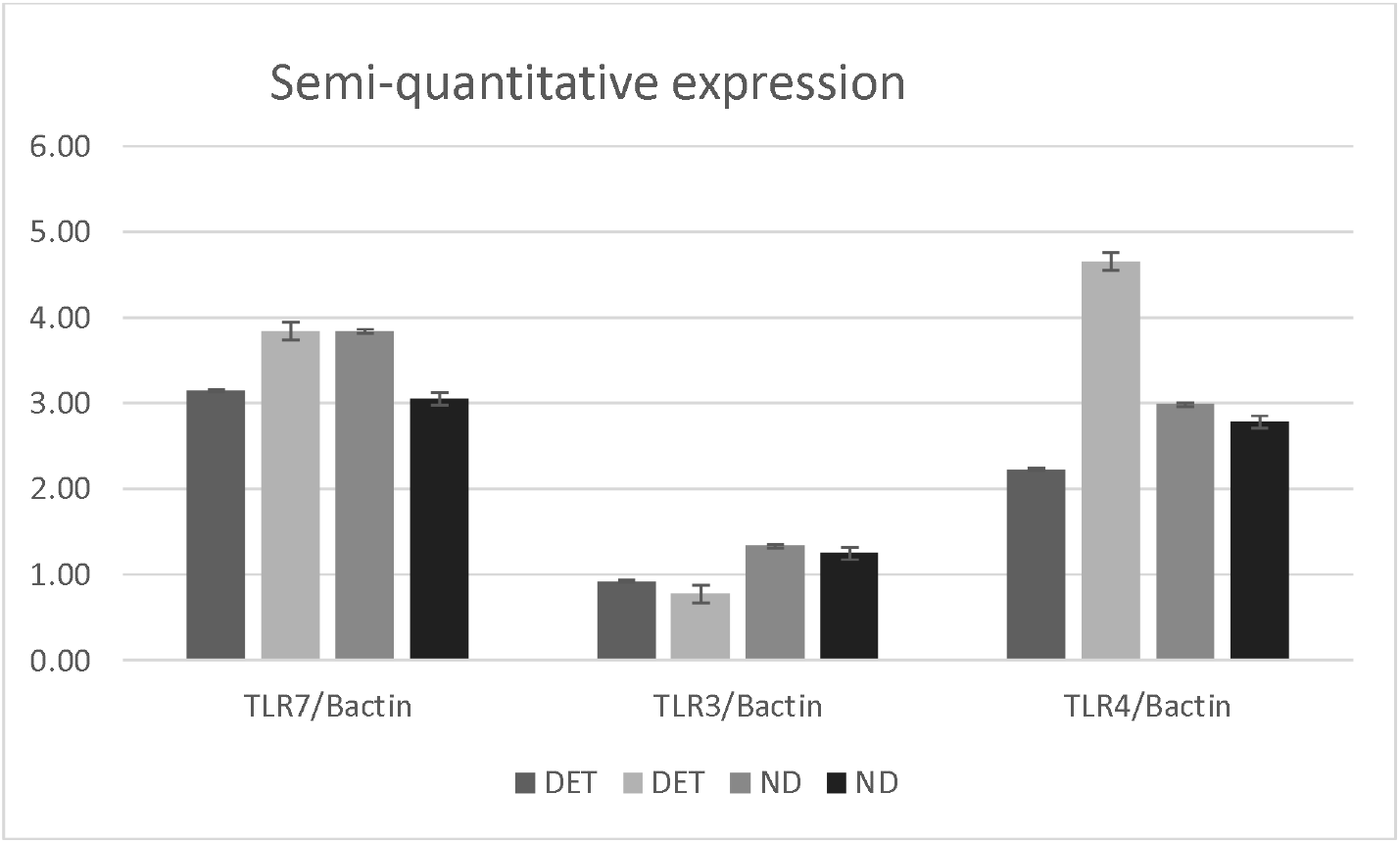
TLRs expression in the 4 studied cohorts.

On the other hand, TLR4 showed a significant higher expression in all patients, DET or ND with severe symptoms (Sperman’s *r*_*s*_ = 0.15054, *p* (2-tailed) = 0.00925). The comparison of our 4 cohorts revealed that TLR4 expression was two-fold increase in severe patients with COVID-19 infection (T-test p Value <0.001). When we accounted only for ND COVID-19 patients and SEVERITY we evidenced a significant positive correlation for TLR4 (Spearman’s and Point-Biserial Correlations; *r*_*s*_ = 0.26329, *p* (2-tailed) = 0.00118).

When compared SEVERITY of symptoms and gene expression by a Spearman’s Correlation Coefficient there was significant positive correlation between severe symptomatology and death and TLR7 expression in both COVID-19 infected and non-infected patients (rs=0.117 p Value=0.042). The Correlation Coefficient showed that TLR7 expression was significant positive in those ND patients with severe symptomatology (*r*_*s*_ = 0.23909, *p* (2-tailed) = 0.00311).

We also showed analysis for these genes expression by SEX and AGE of patients. In DET samples the Game-Howell’s test gave significant differences between TLR7, TLR3 and TLR4 expression by SEX. For the total samples, women showed a significant severe symptomatology independent of being infected by COVID-19 or not (unpaired T-Test pValue= 0.04). Whereas any evident difference in the number between women or men infected or not with COVID-19 (p Value>0.9) was not detected.

DET and ND have the same patient’s age distribution and a significant small negative difference of expression for TLR3 and TLR7 was found, whereas for TLR4 we evidenced a significant negative difference in expression between young adults, adults and older ones. The older adults yielded significant lower expression, and the adults bear highest expression for TLR4 in these cohorts (p-Value <0.002; *r*_*s*_ = -0.11917, *p* (2-tailed) = 0.03979).

Results of the Pearson correlation indicated that there is a significant very small negative relationship between SEVERITY and AGE, (*r* (300) = 0.126, *p* = 0.029), in both non detected and detected COVID-19 patients, meaning that younger patients of this cohort would develop high severity of symptoms in both groups DET and ND.

## DISCUSSION

Previous work of Menezes et al. (2021) in 79 patients with severe COVID-19 on admission, according to World Health Organization (WHO) classification, were divided into two groups: patients who needed mechanical ventilation and/or deceased (SEVERE, n = 50) and patients who used supplementary oxygen but not mechanical ventilation and survived (MILD, n = 29). A control group (CONTROL, n = 17) was enrolled too. Instead of characterizing gene expression on nasopharyngeal tissue, they use the peripheral blood for gene expression (mRNA) of Toll-like receptors (TLRs) 3, 4, 7, 8, and 9, as well as other immune response genes (RIGI, NLRP3, IFN-α, IFN-β, IFN-γ, IFN-λ, pro-interleukin(IL)-1β (pro-IL-1β), and IL-18 determined on admission, between 5–9 days, and between 10–15 days. Circulating cytokines in plasma were also measured. When compared to the COVID-19 MILD group, the COVID-19 SEVERE group had lower expression of TLR3 and overexpression of TLR4 (Menezes et al, 2021). A difference with our work is that we informed the expression level of 3 TLRs at the moment of infection determination using nasopharyngeal samples from a larger cohort of different severity levels, ages and sex. Like Menezes et al. (2021) we confirmed that TLR4 is 2-fold increase in expression in a much bigger cohort of COVID-19 positive patients with severe symptomatology.

Other authors, such as Bagheri-Hosseinabadi et al (2022) used the same type of biology samples as in our study, epithelial cells from nasopharyngeal swab (90 COVID-19 patients and 50 controls with similar symptomatology as COVID-19 detected patients). Samples classified into those without symptoms, with symptoms but not hospitalized, and with symptoms and hospitalized. The difference with our work is that they showed that the transcript levels of TLR3, TLR7, TLR8, and TLR9 were over expressed in the COVID-19 patients with clinical symptoms needing hospitalization, so this sampling could be correlated to our COVID-19 severe symptomatic patients as well as in those with low clinical symptoms without needing for hospitalization compared to their controls. Brieftly, on the contrary to that work, we did not find differences between the expression of TLR3, TLR4 and TLR7 in those hospitalized non COVID-19 (n=8) or COVID-19 patients (n=5) (data not showed in results) furthermore, no differences in the expression for TLR3 and TLR7 on COVID-19 patients versus those without this infection. They conclude that upregulation of TLRs was associated with clinical presentations of severity of symptoms. For them in that sample of 90 patients and 50 controls the modulation of TLR3, TLR7, TLR8, TLR9 in the epithelial cells of COVID-19 cases may estimate the disease severity and requirement for hospitalization. However, our data may counteract their bias as our cohort size is bigger of this previous study, as we used much larger cohorts for COVID-19 infected or non-infected patients in which we found that TLR4 and TLR7 correlation with severity was significant positive in both cohort, ND or DET patient. When SEVERITY is taken in account TLR4 maintained or support previous results of the higher level of expression in COVID-19 patients with high or severe symptomatology and hospitalization, so TLR4 expression is a factor of unfavorable outcome for COVID-19 infection (Bagheri-Hosseinabadi et al, 2022). For TLR3, the higher expression correlates positively with severe symptoms in those patients without SARS Cov2 infection rejecting previous results such as those from Menezes et al (2021).

On average, all detected patients expressed these genes in a same manner as non-detected patients with severe symptoms, being TLR4 and TLR7 the significant higher expressed genes in all infected COVID-19 patients with severe symptoms. TLR3 and TLR7 also showed a positive correlated expression with those ND patients with low or mild symptomatology. Therefore, we confirmed that the immune response to TLR4 and TLR7 genes expression can be associated with harm and not with protection to the host due to persistent inflammation and tissue destruction in the pathogenesis of COVID□19 and principally, to the other severe respiratory diseases. We suggest that effort must be done to focus therapies on TLR4 and TLR7 expression in inflammatory and severe respiratory diseases with COVID-19 infection. At the same time, the role of TLR7 in different respiratory pathologies need to be evaluated as a probable mitigator of symptoms in other respiratory viral infections whereas TLR4 high expression could be focus as a marker or cause of severe symptoms in COVID-19 infections.

In agreement with Onofrio et al (2020), we suggest the TLRs participation in the subsequent development of fatal clinical manifestations in severe COVID-19 patients, and essentially in ARDS (acute respiratory distress syndrome), therefore they would be related with a fatal respiratory failure.

Our data suggest that even though our innate immune system with TLRs contributes to the elimination of viruses, the maintenance of inflammation and tissue destruction associated with harm to the host could be linked with TLR4 in COVID-19 patients. We confirmed that the genes TLR3, TLR4 and TLR7 could be involved in the pathogenesis of other respiratory diseases and the severity of different aged patients. We suggest that treatments focused on these genes expression in inflammatory respiratory diseases could be a start point against unfavorable outcomes and severe symptoms development in younger patients. We underline the importance of sex because of severe symptomatology was women presented as the care group.

We suggest to analysed the repositioning in ARDS and severe COVID-19 infected patients antagonist developed as consequence of tumors studies.

## Data Availability

All data produced in the present study are available upon reasonable request to the authors

## Acknowledgements

The authors would like to thanks for samples collection and storage to the Clinical Laboratory of San Jose Hospital, specially Alejandra Chemez BSc biochemistry; Jose Luis Favant PhD Bch and Dr Carina Reh and Vazquez Sonia, authorities from the Ministry of Health Province of Entre Rios, for their support in optimizing Neo Kit Plus for the rapid genetic testing by isothermal technique of COVID-19.

